# Assessment of Minidock MTB for the diagnosis of tuberculosis from sputum in patients presenting to health facilities in Indonesia

**DOI:** 10.64898/2026.04.22.26351245

**Authors:** Sri Hartati, Raspati C Koesoemadinata, Katrina Sharples, Susan McAllister, Lidya Chaidir, Irmawanty Setiaputri, Emmanuel, Reinout van Crevel, Stephen M Graham, Philip C Hill, Bachti Alisjahbana

**Affiliations:** Research Center for Care and Control of Infectious Disease, Universitas Padjadjaran, Bandung, Indonesia; Faculty of Medicine, Universitas Pendidikan Indonesia, Bandung, Indonesia; Centre for International Health, Division of Health Sciences, University of Otago, Dunedin, New Zealand; Department of Medicine, Faculty of Medicine – Dunedin, University of Otago, Dunedin, New Zealand; Department of Biomedical Sciences, Faculty of Medicine, Universitas Padjadjaran, Bandung, Indonesia; Dr. H. A. Rotinsulu Lung Hospital, Bandung, Indonesia; Dr. H. A. Rotinsulu Cibadak Primary Clinic, Bandung, Indonesia; Department of Internal Medicine, Radboud University Nijmegen Medical Centre, Nijmegen, The Netherlands; Blizard Institute, Faculty of Medicine and Dentistry, Queen Mary University of London, UK; The University of Melbourne Department of Paediatrics and Murdoch Children’s Research Institute, Royal Children’s Hospital, Melbourne, Australia; Department of Internal Medicine, Faculty of Medicine, Universitas Padjadjaran/Hasan Sadikin General Hospital, Bandung, Indonesia

**Author notes:** Equal contribution as first author (Sri Hartati, Raspati C Koesoemadinata). **Corresponding author:** Bachti Alisjahbana, Department of Internal Medicine, Faculty of Medicine, Universitas Padjadjaran/Hasan Sadikin General Hospital, Bandung, Indonesia., Bandung City, Indonesia.

**Keywords:** tuberculosis, diagnostic, MiniDock, sputum swab

## Abstract

**Background:** Access to tuberculosis (TB) diagnostics remains limited in high-burden countries, partly due to centralised and complex testing. We evaluated MiniDock MTB, a low-complexity near point-of-care (nPOC) assay, in sputum for diagnostic accuracy and agreement with Xpert MTB/RIF Ultra (Xpert).

**Methods:** From September 2024 to April 2025, presumptive pulmonary TB cases aged >28 days were consecutively enrolled at 15 community health centres, a lung clinic, and a lung hospital in Bandung, Indonesia. Sputum was tested with MiniDock MTB on sputum swab, Xpert, and liquid culture. We assessed diagnostic accuracy against microbiological and composite reference standards (CRS) and agreement with Xpert.

**Results:** From 3051 individuals screened, 671 were eligible and included; 533 were adults (aged ≥15 years), 138 were children. Overall, 126 were Xpert-positive and 132 culture-positive. In adults, MiniDock MTB sensitivity was 86.2% (100/116; 95% CI 78.8 – 91.3) and 55.8% (110/197; 95% CI 48.9 – 62.6) against liquid culture and CRS, respectively; small numbers of positive results precluded estimation in children. Specificity ranged from 96.8% to 98.8%. Overall agreement between MiniDock MTB and Xpert (excluding trace positive) was 94.8% (95% CI 92.6 – 96.3; K = 0.84). Positive percent agreement was 82.8% (95% CI 75.1 – 88.4) and 25% (95% CI 4.6 – 69.9) in adults and children, respectively, and reduced with lower bacillary burden (p = 0.004).

**Conclusions:** Sensitivity of MiniDock MTB in sputum against liquid culture exceeded the WHO threshold for a sputum-based nPOC TB test in adults. There was high agreement with Xpert but reduced sensitivity in low-bacillary burden TB disease.

## BACKGROUND

It is estimated that there were 10.7 million incident tuberculosis (TB) cases and 1.23 million deaths in 2024.^1^ However, only 8.3 million cases were diagnosed, and of these, just 4.5 million were identified using a molecular World Health Organization (WHO)-recommended rapid diagnostic tests such as Xpert MTB/RIF Ultra (Xpert). Of 30 high TB burden countries, only eight reported >50% coverage of these tests across diagnostic sites.^1^ These gaps are attributed to cost and supply constraints, infrastructure limitations, and operational complexity,^2–4^ which remain major barriers to TB diagnosis. The diagnostic gap is higher in children, especially in young children (<5 years), who account for nearly 80% of the more than 200,000 annual deaths from undiagnosed paediatric TB.^1^

A new generation of diagnostic platforms for infectious diseases emerged during the COVID-19 pandemic.^5^ Novel molecular TB tests on sputum and non-sputum samples offer lower cost, minimal infrastructure requirements, and the ability to test near the point-of-care (nPOC). One such assay, Minidock MTB (Guangzhou Pluslife Biotech, China), can be used with tongue and sputum swab samples.^6–8^ Results from multi-centre studies suggest that this assay achieves sensitivity and specificity exceeding the WHO target product profile (TPP) thresholds for an nPOC TB diagnostic test, but test sensitivity may be lower than Xpert on sputum.^6–9^ There are limited data on Minidock MTB’s accuracy and potential to replace Xpert as an initial diagnostic test in presumptive TB patients in high-burden settings, and none from Indonesia, which holds 10% of the global TB burden.^1^

Therefore, the EVIDENT Indonesia (Evaluation and Demonstration of New Tuberculosis Diagnostics for Indonesia) research platform aims to evaluate accuracy, feasibility, acceptability, cost-effectiveness and implementation of novel TB diagnostic assays in people of all ages with presumptive TB in Indonesia. In this study, we assessed the accuracy of the MiniDock TB test in sputum samples from symptomatic presumptive TB patients using microbiological and composite reference standards. We also assessed agreement with Xpert and whether results varied by bacillary burden, age, gender, or study site.

## METHODS

### Study design

We conducted a prospective study in Bandung, Indonesia, at a tertiary lung hospital, a secondary lung clinic, and 15 Community Health Centres (CHCs) within an established Xpert referral network linked to both sites. We consecutively enrolled patients presenting to these facilities with symptoms suggestive of pulmonary TB. For adults, this was defined as a cough lasting at least two weeks accompanied by at least one additional symptom (fever, night sweats, or unintentional weight loss). Children and adolescents aged 1 month to 17 years were identified as having presumptive pulmonary TB by the doctor in charge following clinical evaluation, including symptoms, contact history and nutritional status. Exclusion criteria included prior TB treatment for more than one week, having received TB preventive therapy within the previous six months, use of antibiotics with anti-TB activity within the past 60 days, a history of TB within the previous two years, or severe illness. Adults unable to produce an adequate sputum sample were also excluded.

### Study procedures

Eligible participants underwent a structured interview to collect demographic and clinical data, along with anthropometric measurements. A Tuberculin Skin Test (TST) was performed routinely in paediatric and, selectively, in adults when indicated. All participants were referred for chest X-ray (CXR), interpreted independently by a radiologist for clinical decision-making. CXRs of children and adolescents were retrospectively reviewed by the study paediatrician. Spontaneous sputum samples were collected up to three samples and characterised macroscopically. The first and second sputum samples were used for Xpert testing. The third sputum sample was used for sputum swab, the remainder was used for the Mycobacteria Growth Indicator Tube (MGIT) culture. If an adequate baseline sample could not be obtained, participants were given sputum pots to collect spontaneous samples over the next three days. Clinicians ordered Xpert testing for participants who could provide sputum, as per national guidelines. Venous blood tests, including HbA1c and HIV, were performed routinely in adults and in children as indicated.

Clinicians made treatment decisions based on clinical presentations and investigations. Participants with uncertain diagnosis were followed up within one month to confirm TB treatment initiation or arrange repeat testing. Among those who initiated TB treatment, treatment response was categorised as: (a) Yes; improvement of TB-related symptoms, (b) No; Lack of improvement, (c) Non-compliance, or (d) Unable to be contacted. Medical records were reviewed to determine alternative diagnoses in those without TB. Data were managed in a Research Electronic Data Capture (REDCap)^10^ database and regularly cross-checked by two researchers.

### Test methods

#### Minidock MTB test (Index test)

Sputum swabs were prepared by swirling a nylon-flocked swab (Copan 502CS01) in sputum sample, then gently wiping it against the container wall to remove excess sputum. Sputum swabs were tested using the MiniDock MTB within 24 hours according to the manufacturer’s instructions (see Supplementary Material). Results were categorised as positive, negative, or invalid/error. Testing was repeated once using the remaining extracted sample if the initial test yielded an invalid or error result.

#### Culture and GeneXpert MTB/RIF Ultra tests (Reference tests)

Sputum specimens were processed for MGIT culture at the lung hospital and Xpert testing at the lung clinic or lung hospital. For culture, sputum was decontaminated with N-acetyl-L-cysteine/sodium hydroxide and tested using the MGIT 960 (BD Microbiology Systems). Participants were classified as culture-positive if *Mycobacterium tuberculosis* (MTB) was identified. When the initial culture yielded nontuberculous mycobacteria (NTM), a second culture from the remaining decontaminated sample was performed to determine the final classification. NTM-positive samples were considered to have no positive MTB result. Xpert testing was performed by mixing sputum with the sample reagent (2:1) and adding 2.0 mL to the cartridge for analysis. The Xpert assay readout reports result as invalid, error, MTB not detected (Xpert negative), or MTB detected (Xpert positive). Semi-quantitative results for Xpert positive were reported as trace, very low, low, medium, or high. For patients referred from other health centres with Xpert positive but lacking semi-quantitative results, results were classified as non-trace if reported as MTB detected, Rifampicin resistance (RR) not detected, or MTB detected, RR detected. The result is categorised as trace if reported as MTB detected, RR indeterminate.^11^ BD MAX testing was performed instead of Xpert in some participants with presumptive drug-resistant TB per national guidelines.

Both index and reference tests were conducted by laboratory personnel who were blinded to clinical data and index or reference test results.

### Statistical analysis

To estimate the diagnostic accuracy of sputum swab MiniDock, the protocol prespecified three reference standards: (1) a microbiological reference standard (MRS) based on culture or Xpert, (2) an extended MRS (EMRS) based on culture, Xpert or additional clinically indicated testing (Any additional mycobacterial culture or Xpert from other respiratory and/ or non-respiratory samples), and (3) a composite reference standard (CRS). Before analysis, these definitions were modified to better align with the WHO TPP for TB diagnostics, as follows.

#### Reference standards for ≥15 age group

Reference standards used were: i) Strict MRS (SMRS), defined TB-positive as MTB culture-positive, and otherwise TB-negative; ii) EMRS, defined TB-positive as MTB culture-positive or Xpert-positive, otherwise TB-negative; iii) CRS, defined TB-positive as either Definite or Probable TB, and otherwise TB-negative. In a sensitivity analysis, TB-positive cases included Definite, Probable and Possible TB. To derive the CRS, patients were classified as: a) Definite TB: Microbiological confirmation obtained (MTB confirmed in culture, or Xpert – excluding trace, or BD MAX), b) Probable TB: Microbiological confirmation not obtained, but CXR suggestive of active TB and initiation of TB treatment; including Xpert positive trace, c) Possible TB: No microbiological confirmation, with either CXR suggestive of active TB or initiation of TB treatment, d) Not TB: Negative on at least one of culture, Xpert, BD MAX, and CXR, with others not done, no TB treatment (Figure 1 and Supplementary Table S1).

**Figure 1.**
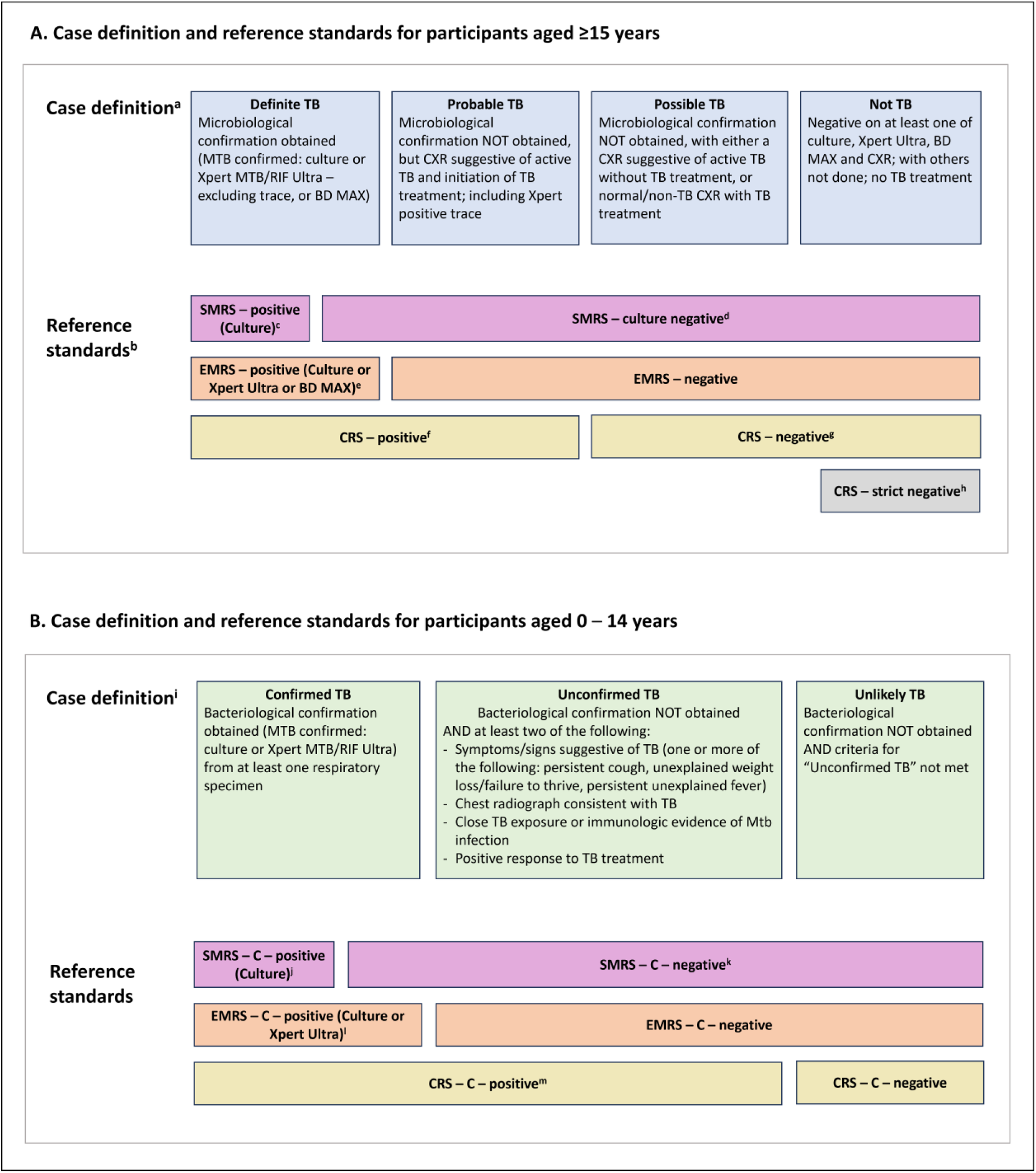
Clinical case definitions and reference standards for participants. (A) ^a^As per Table S1; ^b^Reference standards as per supplementary Table S1; ^c^SMRS = Strict microbiological reference standard; ^d^Criteria for age ≥15 is negative culture; ^e^EMRS = Extended microbiological reference standard; ^f^CRS = Composite reference standard; ^g^Those who are not positive according to CRS; ^h^Those negative on at least two of culture, Xpert and CXR, with the third not done, and not treated for TB. (B) ^i^As per the National Institute of Health (NIH) categorisation as defined by Graham et al for children aged 0–14 years; ^j^SMRS – C = SMRS – Children; ^k^Criteria for age 0 – 14 is the absence of positive culture; ^l^EMRS – C = EMRS – Children; ^m^CRS – C = Composite reference standard – Children.

#### Reference standards for 0 – 14 age group

Reference standards used were: (i) SMRS – Children (SMRS – C) defined TB-positive as MTB culture-positive, and TB-negative otherwise; (ii) EMRS – C defined TB-positive as culture-positive or Xpert-positive, and TB-negative otherwise; (iii) CRS – C defined TB-positive as Confirmed TB or Unconfirmed TB, and TB-negative otherwise (Figure 1). Confirmed, Unconfirmed, and Unlikely TB were classified according to the National Institutes of Health (NIH) paediatric TB case definitions.^12^

Across all analyses, estimates of sensitivity, specificity, positive predictive value (PPV), and negative predictive value (NPV) were calculated, with corresponding 95% confidence intervals (CI) computed using Wilson’s score method. To determine the performance of the sputum swab MiniDock test in participants without TB (CRS strict negative), we calculated the proportion negative and 95% CI among participants who were not treated for TB and had negative results on two of three tests – culture, Xpert or BD MAX, and CXR – while the third was not performed.

Agreement between the sputum swab MiniDock and Xpert on sputum was assessed both excluding and including trace results. Secondary analyses assessed agreement stratified by (i) bacillary burden, using the semi-quantitative categories of Xpert on sputum, (ii) age group (0 – 14 and ≥15 years), (iii) sex, and (iv) study site. Agreement was assessed as (i) the proportion of sputum Xpert-positives who were sputum swab MiniDock-positive (positive percent agreement (PPA)), (ii) the proportion of sputum Xpert-negatives who were sputum swab MiniDock-negative (negative percent agreement (NPA)) and (iii) the overall agreement, with 95% CI computed using Wilson’s score method. Differences in PPA and NPA across subgroup levels were compared using the Chi-square test of association for age, sex and study site, and for trend for bacillary burden. P-values less than 0.05 were considered significant. Level of agreement was evaluated using Cohen’s Kappa coefficient (K) and 95% CI. Xpert-positive results lacking semi-quantitative categories were classified as trace or non-trace according to their Xpert readout (as defined in the Methods); non-trace results were included in the primary analysis but excluded from stratified analyses by bacillary burden. Invalid/error results were excluded from agreement and diagnostic accuracy analysis.

### Sample size

Sample size for the EVIDENT studies was primarily determined by the precision required for estimating the proportion of individuals testing positive by an index test among those who were positive by reference standards. We assumed sensitivities of 70–80% and a TB prevalence of 15%.^9^ To obtain a CI of width of ±5% around the sensitivity estimate, we aimed to enrol approximately 600 participants with presumptive pulmonary TB.

Data were analysed using Stata 18.0 (StataCorp LLC, College Station, TX, USA)^13^ and R 4.5.0 (R Foundation, Vienna).^14^ This study is reported according to the Standards for Reporting of Diagnostic Accuracy Studies (STARD) guidelines (Supplementary Table S7).^15^

### Ethics

The study protocol was approved by the Health Research Ethics Committee of Universitas Padjadjaran (Ref. No. 616/UN6.KEP/EC/2024). Written informed consent was obtained from all adult participants (≥18 years) and from the parents or legal guardians of participants aged <18 years. Assent was obtained from children aged ≥12 years.

## RESULTS

Between September 2024 and April 2025, 3051 individuals were screened, and 677 eligible participants were enrolled. Six were excluded from the analysis due to a suspected laboratory contamination event (Figure 2). Of the 671 analysed, 20.6% were aged 0 – 14 years and 3.1% were <5 years old, 44.3% were female, 1.3% were diagnosed with HIV, and 15.1% were diagnosed with diabetes (Table 1). Half (49.9%) were recruited from the lung clinic, 28.6% from the hospital and 21.5% from CHCs. Demographic and clinical characteristics varied by site (Supplementary Table S2).

**Figure 2.**
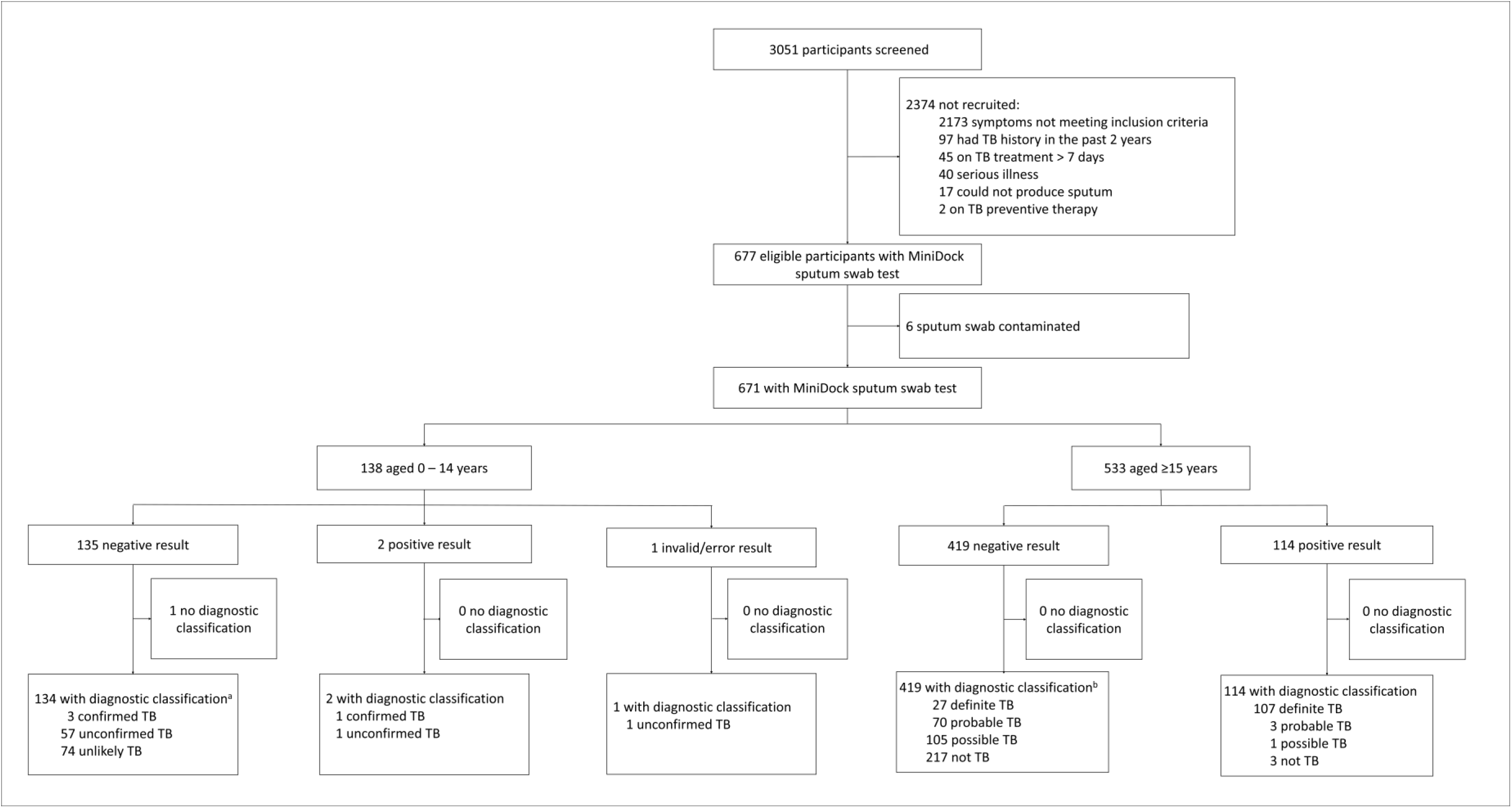
Participant flow diagram for MiniDock sputum swab. ^a^Patients aged 0 – 14 years old were categorised as per the NIH categorisation as defined by Graham et al: Confirmed TB defined as bacteriological confirmation obtained for culture or Xpert; Unconfirmed TB defined as bacteriological confirmation NOT obtained AND at least 2 of the following: symptoms/signs suggestive of TB, CXR consistent of TB, close TB exposure or immunologic evidence of MTB infection, positive response to TB treatment; Unlikely TB defined as bacteriological confirmation NOT obtained AND criteria for “Unconfirmed TB” not met, ^b^Patients aged ≥15 were categorised as follows: Definite TB defined as positive by culture for MTB or by Xpert, excluding trace; Probable TB defined as culture and Xpert negative or not done, but CXR suggestive of active TB and treated for TB; Possible TB defined as (i) CXR suggestive of active TB without microbiological confirmation and no TB treatment initiated, or (ii) CXR normal or non-TB but started on TB treatment; Not TB defined as negative culture, Xpert, and CXR, with no TB treatment.

**Table 1.** Demographic, clinical characteristics and laboratory findings of eligible patients by age group (n=671)

| Characteristic | 0 – 4 years<br>(n=21)<br>n (%) | 5 – 9 years<br>(n=70)<br>n (%) | 10 – 14 years<br>(n=47)<br>n (%) | ≥15 years<br>(n=533)<br>n (%) | Total<br>(n=671)<br>n (%) |
| --- | --- | --- | --- | --- | --- |
| Female | 15 (71.4) | 38 (54.3) | 25 (53.2) | 219 (41.1) | 297 (44.3) |
| Hospitalisation at enrolment | 1 (4.8) | 1 (1.4) | 2 (4.3) | 135 (25.3) | 139 (20.7) |
| **Symptoms** |  |  |  |  |  |
| Cough |  |  |  |  |  |
| Persistent (≥14 days) | 14 (66.7) | 54 (77.1) | 34 (72.3) | 521 (97.8) | 623 (92.9) |
| Acute (<14 days) | 4 (19.0) | 6 (8.6) | 5 (10.6) | 6 (1.1)<sup>a</sup> | 21 (3.1) |
| None | 3 (14.3) | 10 (14.3) | 8 (17.0) | 6 (1.1)<sup>a</sup> | 27 (4.0) |
| Additional symptoms<sup>b</sup> |  |  |  |  |  |
| Two or more | 15 (71.4) | 40 (57.1) | 30 (63.8) | 447 (83.9) | 532 (79.3) |
| One | 6 (28.6) | 24 (34.3) | 15 (31.9) | 79 (14.8) | 124 (18.5) |
| None | 0 (0.0) | 6 (8.6) | 2 (4.3) | 7 (1.3) | 15 (2.2) |
| **TB risk factors** |  |  |  |  |  |
| Underweight<sup>c</sup> | 1 (4.8) | 18 (25.7) | 5 (10.6) | 186 (34.9) | 210 (31.3) |
| Smoking – yes | 0 (0.0) | 0 (0.0) | 0 (0.0) | 179 (33.6) | 179 (26.7) |
| Reported TB contact | 11 (52.4) | 28 (40.0) | 23 (48.9) | 98 (18.4) | 160 (23.8) |
| Previous TB treatment (≥2 years ago) | 0 (0.0) | 6 (8.6) | 5 (10.6) | 151 (28.3) | 162 (24.1) |
| HIV diagnosis<sup>d</sup> |  |  |  |  |  |
| Yes | 0 (0.0) | 0 (0.0) | 0 (0.0) | 6 (1.4) | 6 (1.3) |
| No | 0 (0.0) | 1 (100) | 3 (100) | 440 (98.6) | 444 (98.7) |
| Unknown<sup>e</sup> | 21 | 69 | 44 | 87 | 221 |
| Diabetes Mellitus diagnosis<sup>f</sup> |  |  |  |  |  |
| Yes | 0 (0.0) | 0 (0.0) | 1 (9.1) | 77 (15.8) | 78 (15.1) |
| No | 0 (0.0) | 18 (100) | 10 (90.9) | 412 (84.2) | 440 (84.9) |
| Unknown | 21 | 52 | 36 | 44 | 153 |
| **Clinical investigations** |  |  |  |  |  |
| Tuberculin Skin Test (TST) |  |  |  |  |  |
| TST positive | 10 (50.0) | 33 (48.5) | 22 (59.5) | 20 (74.1) | 85 (55.9) |
| TST negative | 10 (50.0) | 35 (51.5) | 15 (40.5) | 7 (25.9) | 67 (44.1) |
| Not done | 1 | 2 | 10 | 506 | 519 |
| Chest X-ray |  |  |  |  |  |
| TB-specific abnormality | 12 (63.1) | 52 (75.4) | 27 (57.4) | 296 (56.3) | 387 (58.7) |
| Non-TB abnormality | 6 (31.6) | 14 (20.3) | 13 (27.7) | 162 (30.9) | 195 (29.5) |
| Normal | 1 (5.3) | 3 (4.3) | 7 (14.9) | 67 (12.8) | 78 (11.8) |
| Not readable | 1 | 0 | 0 | 0 | 1 |
| Not done | 1 | 1 | 0 | 8 | 10 |
| **Laboratory results** |  |  |  |  |  |
| Molecular test performed |  |  |  |  |  |
| Xpert | 1 (4.8) | 38 (54.3) | 39 (83.0) | 508 (95.3) | 586 (87.3) |
| BD MAX | 0 (0.0) | 0 (0.0) | 0 (0.0) | 18 (3.4) | 18 (2.7) |
| Not done | 0 (0.0) | 0 (0.0) | 0 (0.0) | 7 (1.3) | 7 (1.0) |
| No sputum | 20 (95.2) | 32 (45.7) | 8 (17.0) | 0 (0.0) | 60 (8.9) |
| Xpert result |  |  |  |  |  |
| MTB detected | 0 (0.0) | 0 (0.0) | 4 (10.3) | 134 (26.4) | 138 (23.6) |
| MTB not detected | 1 (100) | 38 (100) | 35 (89.7) | 374 (73.6) | 448 (76.4) |
| Xpert semi-quantitative result |  |  |  |  |  |
| High | 0 (0.0) | 0 (0.0) | 2 (50.0) | 40 (29.9) | 42 (30.4) |
| Medium | 0 (0.0) | 0 (0.0) | 0 (0.0) | 17 (12.7) | 17 (12.3) |
| Low | 0 (0.0) | 0 (0.0) | 2 (50.0) | 44 (32.8) | 46 (33.3) |
| Very low | 0 (0.0) | 0 (0.0) | 0 (0.0) | 11 (8.2) | 11 (8.0) |
| Trace | 0 (0.0) | 0 (0.0) | 0 (0.0) | 12 (8.9) | 12 (8.7) |
| Not recorded | 0 (0.0) | 0 (0.0) | 0 (0.0) | 10 (7.5) | 10 (7.3) |
| BD MAX result |  |  |  |  |  |
| MTB detected | 0 (0.0) | 0 (0.0) | 0 (0.0) | 3 (0.6) | 3 (0.6) |
| MTB not detected | 0 (0.0) | 0 (0.0) | 0 (0.0) | 15 (83.3) | 15 (83.3) |
| Culture results |  |  |  |  |  |
| Positive | 1 (5.0) | 0 (0.0) | 2 (4.3) | 129 (24.2) | 132 (19.8) |
| Negative | 19 (95.0) | 68 (100) | 45 (95.7) | 404 (75.8) | 536 (80.2) |
| Not done<sup>g</sup> | 1 | 2 | 0 | 0 | 3 |
| Culture positive identification |  |  |  |  |  |
| Mycobacterium tuberculosis | 0 (0.0) | 0 (0.0) | 2 (100) | 116 (89.9) | 118 (89.4) |
| Nontuberculous Mycobacterium | 1 (100) | 0 (0.0) | 0 (0.0) | 13 (10.1) | 14 (10.6) |
| **TB Treatment** |  |  |  |  |  |
| Treated for TB |  |  |  |  |  |
| Yes | 14 (66.7) | 39 (55.7) | 25 (53.2) | 218 (40.9) | 296 (44.1) |
| No | 7 (33.3) | 31 (44.3) | 22 (46.8) | 315 (59.1) | 375 (55.9) |
| Treatment response |  |  |  |  |  |
| Yes | 11 (78.6) | 29 (74.4) | 17 (68.0) | 106 (48.6) | 163 (55.1) |
| No | 1 (7.1) | 2 (5.1) | 2 (8.0) | 7 (3.2) | 12 (4.1) |
| Non compliance | 2 (14.3) | 6 (15.4) | 5 (20.0) | 61 (28.0) | 74 (25.0) |
| Unable to be contacted | 0 (0.0) | 2 (5.1) | 1 (4.0) | 44 (20.2) | 47 (15.9) |
| **Diagnostic classification for 0 – 14-year-olds<sup>h</sup>** |  |  |  |  |  |
| Confirmed TB | 0 (0.0) | 0 (0.0) | 4 (8.5) | N/A | N/A |
| Unconfirmed TB | 12 (57.1) | 34 (48.6) | 13 (27.7) | N/A | N/A |
| Unlikely TB | 8 (38.1) | 36 (51.4) | 30 (63.8) | N/A | N/A |
| Unclassifiable | 1 (4.8) | 0 (0.0) | 0 (0.0) | N/A | N/A |
| **Diagnostic classification for 15 years or older<sup>i</sup>** |  |  |  |  |  |
| Definite TB | N/A | N/A | N/A | 134 (25.1) | N/A |
| Probable TB | N/A | N/A | N/A | 73 (13.7) | N/A |
| Possible TB | N/A | N/A | N/A | 106 (19.9) | N/A |
| Not TB | N/A | N/A | N/A | 220 (41.3) | N/A |

Of the 533 participants aged ≥15 years, 525 underwent CXR; 296 (56.4%) had findings consistent with TB. All participants provided sputum samples with a volume of ≥3 mL. Almost half (261; 49.0%) were mucoid and 79 (14.8%) were purulent. All underwent culture testing; 129 (24.2%) tested positive. Among 508 who underwent Xpert testing, 134 (26.4%) tested positive for MTB; 54.0% had low or lower semi-quantitative results. Semi-quantitative results were not recorded for 10 participants. Of these, seven had a readout of MTB detected, Rifampicin resistance (RR) not detected and three had a readout of MTB detected, RR detected. Those 10 patients were classified as non-trace.^11^ Overall, 134 (25.1%) were classified as Definite TB.

Of 138 participants aged 0 – 14 years, 113 were from the lung clinic; 136 underwent CXR; 91 (66.9%) had findings consistent with TB. Sputum was obtained from all; 120 (87.0%) had a volume ≥2 mL and most were salivary. Only 135 cultures could be performed; three were culture-positive. Of 78 Xpert tests ordered by clinicians, four were Xpert-positive, all aged 10 – 14 years. Of 137 with NIH classification, four were Confirmed TB.

Of the 671 MiniDock tests performed, 8 (1.2%) initially yielded invalid or error results. After performing a single repeat per invalid/error test, only one (0.1%) of 671 remained invalid/error. The time to generate negative results was 25 minutes, while the average time to generate positive results was 16.1 minutes (SD 5.1 minutes).

### Estimated accuracy of the sputum swab MiniDock MTB against reference standards

Among participants aged ≥15 years, the sensitivity was 86.2% (100/116; 95% CI 78.8 to 91.3) using SMRS, 79.9% (107/134; 95% CI 72.3 to 85.8) with the EMRS, and 55.8% (110/197; 95% CI 48.9 to 62.6) using the CRS. Sensitivity was slightly reduced for both EMRS and CRS when Xpert trace results were treated as positive. Specificity ranged from 96.8% to 98.8% across reference standards. In TB-negative cases (CRS – strict negative), specificity was 98.6% (218/220; 95% CI 96.1 to 99.5). (Figure 3 and supplementary Table S6). Among patients aged 0 – 14 years, sensitivity was 50% (1/2; 95% CI 9.4 to 90.5) using SMRS – C, 25% (1/4; 95% CI 4.6 to 69.9) with the EMRS – C, and 3.2% (2/62; 95% CI 0.9 to 11.0) with the CRS – C. Specificity ranged from 99.2% to 100%.

**Figure 3.**
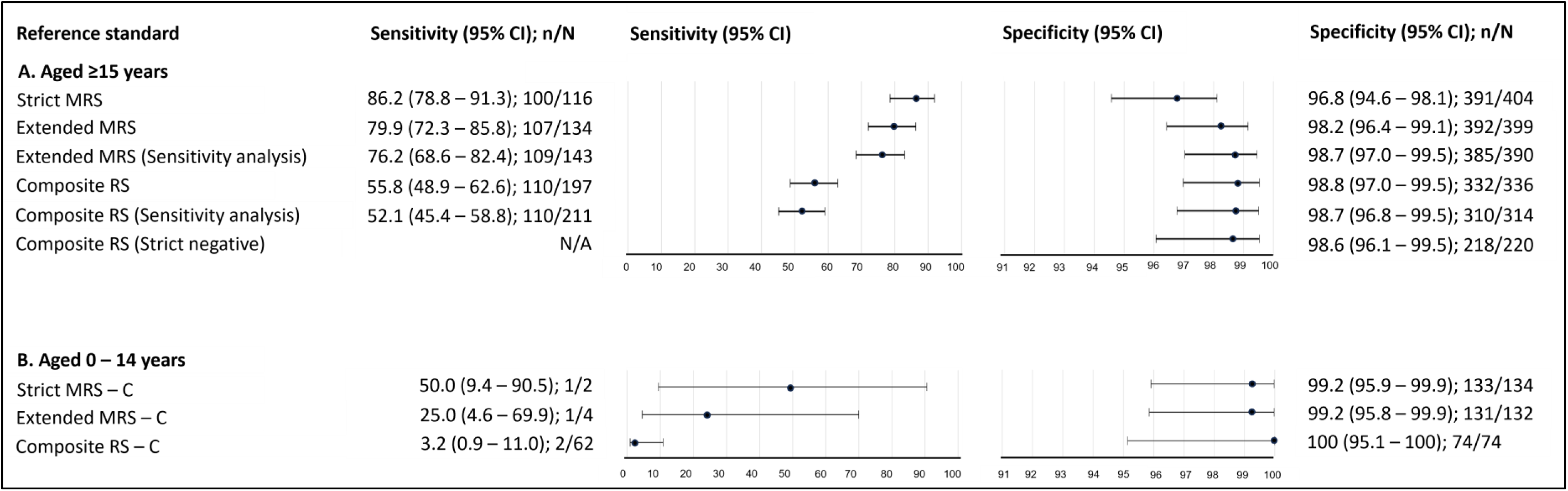
Diagnostic accuracy of MiniDock sputum swab across reference standards stratified by age group. MiniDock sputum swab diagnostic performance assay by reference standard, stratified by age group (0 – 14 years and ≥15 years). Points show point estimates and horizontal bars 95% confidence intervals (Wilson method); n/N indicates the number of true positives or true negatives over the total tested. Reference standards as defined in Figure 1 and supplemental Table S1. (A) Reference standard for patients aged 0 – 14, (B) Reference standard for patients aged ≥15. Abbreviations: MRS, microbiological reference standard; MRS – A, MRS – Adult; RS, Reference standard; Strict MRS defined TB-positive as MTB culture-positive, and TB-negative otherwise; Extended MRS defined TB-positive as culture-positive orXpert-positive, and TB-negative otherwise; Composite RS defined TB-positive as confirmed TB or unconfirmed TB, and TB-negative otherwise; Strict MRS - A defined TB-positive as MTB culture-positive, and otherwise TB-negative; Extended MRS – A, defined TB-positive as MTB culture positive or Xpert positive, otherwise TB-negative; Composite RS – A defined TB-positive as Definite or Probable TB, and otherwise TB-negative; Composite RS – A (Sensitivity analysis) defined TB-positive as Definite, Probable and Possible TB; Composite RS – A (Strict negative) defined TB-negative as i) culture negative, Xpert negative, CXR negative and not treated for TB; or ii) culture negative, either Xpert negative or CXR negative, with the other not done, and not treated for TB.

### Agreement between the MiniDock and Xpert on sputum in the total population

There were 573 participants with both Minidock and Xpert (excluding trace) results; the PPA was 80.9% (102/126; 95% CI, 73.2 to 86.9), while the NPA was 98.7% (441/447; 95% CI, 97.1 to 99.4) (Table 2). Overall agreement was 94.8% (543/573; 95% CI, 92.6 to 96.3). In the sensitivity analysis of 585 participants, including Xpert trace results, the PPA was 76.8% (106/138; 95% CI 69.1 to 83.1) and the overall agreement was 93.5% (547/585; 95% CI, 91.2 to 95.2).

**Table 2.** Agreement between MiniDock and Xpert on sputum.

|  | n/N | Positive percent agreement<br>% (95% CI) | P<br>value<sup>a</sup> | n/N | Negative percent agreement<br>% (95% CI) | P<br>value<sup>a</sup> | n/N | Agreement %<br>(95% CI) | Kappa value<br>(95% CI) |
| --- | --- | --- | --- | --- | --- | --- | --- | --- | --- |
| Overall, excluding trace<sup>b</sup> | 102/126 | 80.9 (73.2 – 86.9) | N/A | 441/447<sup>c</sup> | 98.7 (97.1 – 99.4) | NA | 543/573 | 94.8 (92.6 – 96.3) | 0.84 (0.78 – 0.90) |
| Overall, including trace<sup>d</sup> | 106/138 | 76.8 (69.1 – 83.1) | N/A | 441/447 | 98.7 (97.1 – 99.4) | NA | 547/585 | 93.5 (91.2 – 95.2) | 0.81 (0.75 – 0.87) |
| Stratified analysis of agreement between MiniDock and Xpert on sputum |  |  |  |  |  |  |  |  |  |
| Bacillary burden |  |  |  |  |  |  |  |  |  |
| High | 40/42 | 95.2 (84.2 – 98.7) | 0.004 | N/A | N/A | N/A | N/A | N/A | N/A |
| Medium | 14/17 | 82.4 (59.0 – 93.8) |  |  |  |  |  |  |  |
| Low | 33/46 | 71.7 (57.5 – 82.7) |  |  |  |  |  |  |  |
| Very low | 8/11 | 72.7 (43.4 – 90.3) |  |  |  |  |  |  |  |
| Trace | 4/12 | 33.3 (13.8 – 60.9) |  |  |  |  |  |  |  |
| Excluding trace semiquantitative results |  |  |  |  |  |  |  |  |  |
| Age group |  |  |  |  |  |  |  |  |  |
| 0 – 14 | 1/4 | 25.0 (4.6 – 69.9) | 0.004 | 73/73 | 100 (95.0 – 100) | 0.276 | 74/77 | 96.1 (89.2 – 98.7) | 0.39 (0.15 – 0.92) |
| ≥ 15 | 101/122 | 82.8 (75.1 – 88.4) |  | 368/374 | 98.4 (96.5 – 99.3) |  | 469/496 | 94.6 (92.2 – 96.2) | 0.85 (0.79 – 0.90) |
| Sex |  |  |  |  |  |  |  |  |  |
| Female | 33/47 | 70.2 (56.0 – 81.3) | 0.018 | 191/195 | 97.9 (94.8 – 99.2) | 0.352 | 224/242 | 92.6 (88.5 – 95.2) | 0.74 (0.63 – 0.85) |
| Male | 69/79 | 87.3 (78.2 – 92.9) |  | 250/252 | 99.2 (97.1 – 99.8) |  | 319/331 | 96.4 (93.8 – 97.9) | 0.90 (0.84 – 0.95) |
| Site |  |  |  |  |  |  |  |  |  |
| Lung hospital | 59/73 | 80.8 (70.3 – 88.2) | 0.171 | 103/107 | 96.3 (90.8 – 98.5) | 0.113 | 162/180 | 90.0 (84.7 – 93.6) | 0.79 (0.70 – 0.88) |
| Lung clinic | 27/30 | 90.0 (74.4 – 96.5) |  | 228/230 | 99.1 (96.9 – 99.8) |  | 255/260 | 98.1 (95.6 – 99.2) | 0.90 (0.82 – 0.99) |
| CHC | 16/23 | 69.6 (49.1 – 84.4) |  | 110/110 | 100 (96.6 – 100) |  | 126/133 | 94.7 (89.5 – 97.4) | 0.79 (0.64 – 0.94) |

<sup>a</sup>P value from chi-square test comparing positive percent agreement and negative percent agreement, across age, sex and study site categories, and a Chi-square test for trend for bacillary burden;
<sup>b</sup>Xpert trace semi-quantitative results (n=12) were excluded from both the numerator and the denominator. Xpert with not recorded semi-quantitative results (n=10) were treated as non-trace and included in the analysis; <sup>c</sup>Of the total 448 Xpert negative results, one was not included in the calculation due to MiniDock MTB error/invalid result; <sup>d</sup>Xpert trace semi-quantitative results (n=12) were counted as positive.

Thirty-eight (6.5%) of the 585 participants had discordant results: 32 were Xpert-positive but sputum swab MiniDock-negative, of whom 24 had semi-quantitative results of low or lower (Figure 4A). Six were Xpert-negative but sputum swab MiniDock-positive, of whom one was Possible TB and three were Definite TB cases (Figure 4B).

**Figure 4.**
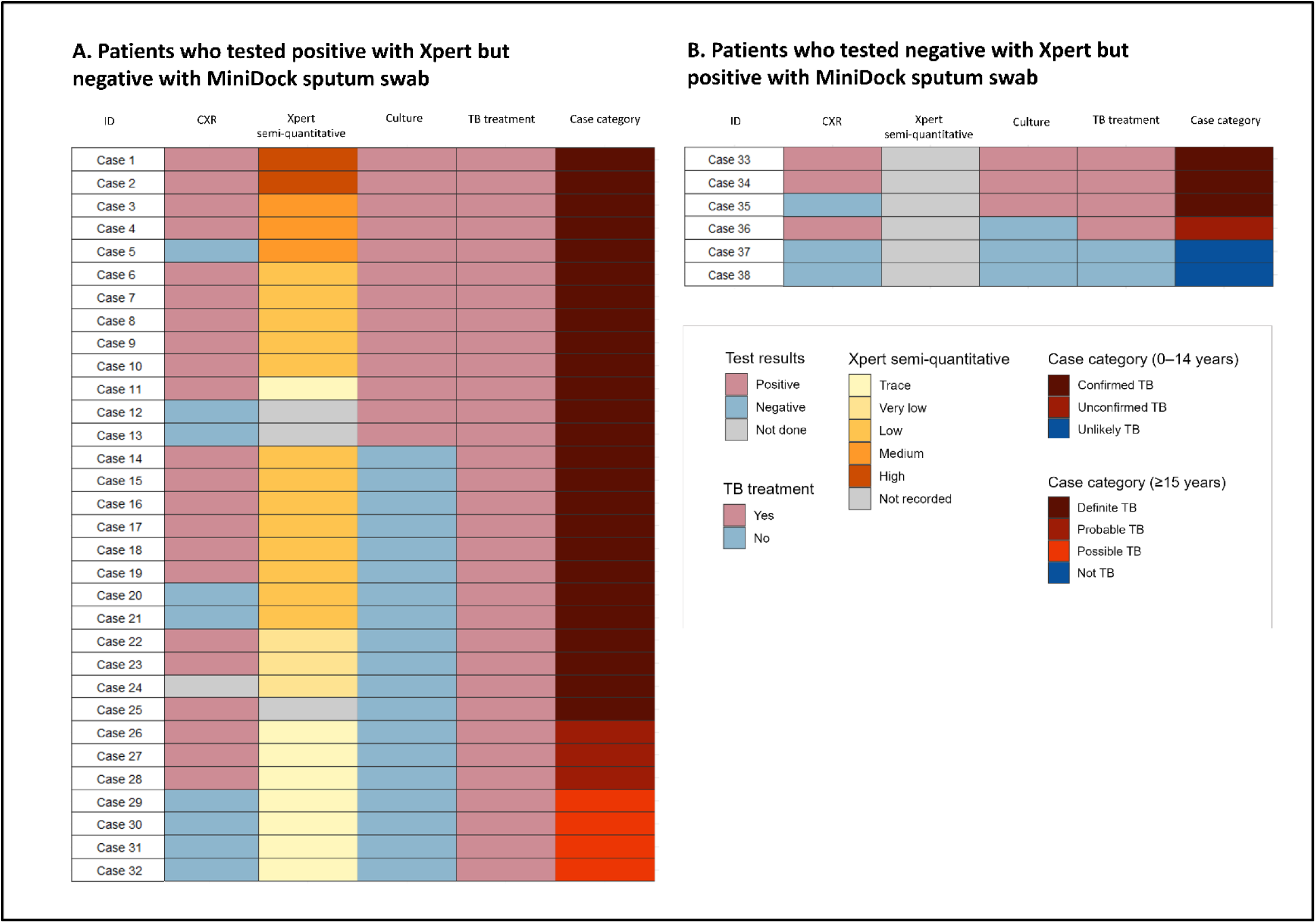
Distribution of discordant patients identified in the agreement analysis between MiniDock sputum swab and Xpert. Description for discordant patients. Each row represents one patient. Each column represents clinical data or tests performed for each patient. (A) Thirty-two patients had Xpert-positive but MiniDock sputum swab-negative results. (B) Six patients had Xpert-negative but MiniDock sputum swab-positive results.

When stratified by bacillary burden, PPA increased with increasing Xpert semiquantitative category (chi-square test for trend: p = 0.004). PPA was lower in children aged 0 – 14 years (25.0%; 95% CI, 4.6 to 69.9%) compared with 82.8% (95% CI, 75.1 to 88.4%) in participants aged ≥15 years (p = 0.004). PPA was lower in females than in males (70.2%; 95% CI, 56.0 to 81.3 vs 87.3%; 95% CI, 78.2 to 92.9; p = 0.018). Across sites, PPA ranged from 69.6% (95% CI, 49.1 to 84.4) at CHCs to 90.0% (95% CI, 71.3 to 88.2) at the lung clinic, but the differences were not statistically significant (p = 0.171). NPA was consistently high across all categories, ranging from 96.3% to 100% (Table 2).

## DISCUSSION

In individuals with presumptive pulmonary TB in urban clinics in Indonesia, we evaluated the accuracy of MiniDock on sputum relative to TB reference standards and agreement with Xpert. In adults, against the SMRS, the sensitivity of 86.2% met the minimum WHO TPP threshold for sputum-based nPOC diagnostics (≥85%), while the specificity of 96.8% did not meet the ≥98% threshold. When evaluated against the EMRS and CRS, specificity was 98.2% and 98.8%, respectively, meeting the ≥98% threshold.^9^ Overall agreement with Xpert was high (94.8%) with a PPA of 82.8% in those aged ≥15 years. PPA was higher in those aged ≥15 years than in children, and in those with higher versus lower Xpert semiquantitative results.

A positive culture, defined as SMRS in our study, is widely used as a reference standard for evaluating TB diagnostics and informing WHO TPP thresholds. Using this reference standard, the sputum swab MiniDock demonstrated sensitivity exceeding the WHO TPP thresholds for sputum-based nPOC tests in adults^9^ and showed comparable sensitivity to a previous Cameroon health-facility study (86.4%),^7^ although slightly lower than the earlier multi-country evaluation (89.9%).^6^ However, SMRS may exclude some patients with TB disease, while CRS may include individuals without TB. In addition, human interpretation of CXR findings may vary by reader expertise, leading to inter- and intra-reader variability. These may have affected our estimates of diagnostic accuracy. Despite this, specificity remained high across age groups and all reference standards.

Our PPA was lower than that reported in the Cameroon health-facility study (86%)^7^ and that observed in the multi-country evaluation (96.9%).^6^ One possible explanation is a difference in bacillary burden distribution, as the performance of MiniDock may vary across populations with differing disease severity.^16,17^ Relative to both studies, our study population had a higher proportion of low, very low, or trace semi-quantitative results (54% compared to 32.1% in the Cameroon study and 39.7% in the multi-country evaluation), although semi-quantitative results were not recorded for 10 patients in our study. A Chinese study reported an even higher PPA (98.2%); however, it is not directly comparable as MiniDock was evaluated against Xpert MTB/RIF rather than Xpert MTB/RIF Ultra.^18^ The stratified analyses support the influence of bacillary burden on the performance of the sputum swab MiniDock. The lower PPA observed in children than in adults in our study likely reflects the paucibacillary nature of TB in children, small sample sizes, and difficulties in obtaining good-quality sputum samples.

A small proportion of samples showed discordant results between Xpert and MiniDock, most commonly when Xpert-positive/MiniDock-negative. These discordant cases were largely associated with low or lower semi-quantitative Xpert grades, suggesting reduced sensitivity at lower bacillary loads. Of the fewer (n=6) Xpert-negative/MiniDock-positive discordant pairs, four had features of clinical or bacteriological TB, suggesting that these did have TB.

When MiniDock is used as a replacement for Xpert, clinical algorithms need to be carefully constructed. TB treatment might be considered when clinical findings and CXR suggest active TB, despite a negative sputum-swab MiniDock. In the discordant pairs in this study, over two-thirds (68.8%; 22/32) of Xpert-positive/MiniDock-negative cases had a CXR abnormality suggestive of active TB. CXR is frequently integrated into the TB diagnostic algorithm.^19^ It has been shown to detect the majority of bacteriologically confirmed TB cases in adults, with an estimated sensitivity of 84.8%.^20^ If CXR is unavailable, a repeat MiniDock test after a short period of follow-up by a clinician may be appropriate. Further studies in Indonesia are assessing the yield from and optimal timing of a second test.

Only two children had a positive sputum swab MiniDock, suggesting limited diagnostic yield in this age group. Of 138 children who had a sputum swab MiniDock, only 56.5% had an available Xpert result, mainly due to the inability to obtain an adequate sputum sample, and no child <10 years had confirmed TB. Collection of adequate sputum and paucibacillary disease are recognised challenges for confirmation of TB in children.^21^ The NIH diagnostic classification for children was appropriate for use in this study, but considerable uncertainty remains as to how many children with TB are represented in the Unconfirmed or Unlikely categories.

Despite lower sensitivity in people with low-bacillary-burden disease, the overall performance characteristics of Minidock are favourable. It offers several advantages over Xpert, including simplified workflows, rapid turnaround time, minimal infrastructure requirements, and much lower cost. The low error rate of the test compared to the WHO threshold may indicate its robustness. Using a battery helps mitigate invalid/error results due to a power outage. The sputum swab MiniDock is a reliable and low-cost alternative for diagnosing TB in adults in health-facility settings.

Our study had some limitations. Adequate sputum specimens were not obtained from children. Further assessment of MiniDock in children requires more invasive procedures that optimise diagnostic confirmation, such as sputum induction or gastric lavage.^22^ Participants were recruited from the hospital, clinic and CHC settings. Further evaluation is needed in broader populations, including asymptomatic individuals, to understand Minidock’s utility in community-based screening and its potential public health impact. Despite these limitations, a key strength of our study is the inclusion of culture, Xpert, CXR, and MiniDock, which enabled participants to undergo both culture and CRS assessment and provided an integrated understanding to inform future implementation.

## CONCLUSION

MiniDock MTB demonstrated sensitivity that exceeded the WHO threshold for a sputum-based nPOC TB test in adults. There was high agreement with Xpert performed on sputum in the healthcare setting, but reduced sensitivity in low-bacillary burden disease.

## Data Availability

All data produced in the present study are available upon reasonable request to the authors

## Acknowledgements

We thank all study participants and their families for their participation. We thank staff of Bandung District Health Office, Dr. H. A. Rotinsulu Lung Hospital, Dr. H. A. Rotinsulu Cibadak Primary Clinic, Puskesmas Ahmad Yani, Puskesmas Astana Anyar, Puskesmas Balai Kota, Puskesmas Cigadung, Puskesmas Cijagra Baru, Puskesmas Cijagra Lama, Puskesmas Cipaku, Puskesmas Ciumbuleuit, Puskesmas Gumuruh, Puskesmas Lio Genteng, Puskesmas Neglasari, Puskesmas Pagarsih, Puskesmas Pelindung Hewan, Puskesmas Suryalaya and Puskesmas Tamblong for their support and participation in the study. We thank Amy Steadman and Jakko van Ingen for their support with study monitoring. We also thank Puneet Dewan and Emily A Kendall for providing feedback during manuscript development. MiniDock reagents were donated by Pluslife.

## Data availability statement

Data are available upon reasonable request.

