## Supplementary material for "Assessment of Minidock MTB for the diagnosis of tuberculosis from sputum in patients presenting to health facilities in Indonesia"

### SUPPLEMENTAL MATERIALS

### 1. Supplemental Methods

#### Methods M1: MiniDock MTB testing procedure in detail

Sputum swabs were collected from sputum samples by swirling a nylon flocked swab (Copan 502CS01) 10 times in the samples for 15 seconds, followed by wiping the swab against the inner wall of the sputum container. The remaining sputum was used for culture and Xpert testing. Immediately after collection, each swab was placed into a tube, sealed, and placed into a leak-proof bag. The samples were then stored in a cool box with ice (2-8°C) and transported to the research laboratory for further processing. At the laboratory, the sputum swab was refrigerated (2-8°C) and tested in order of arrival within 24 hours. For testing, a swab was inserted into a tube containing buffer solution, swirled, and then lysed for 5 minutes. The lysate was subsequently transferred to a reagent card for analysis using the MiniDock MTB platform, following the manufacturer’s instructions for use (IFU).

### 2. Supplemental Figures

#### Figure S1: Operational workflow for MiniDock MTB

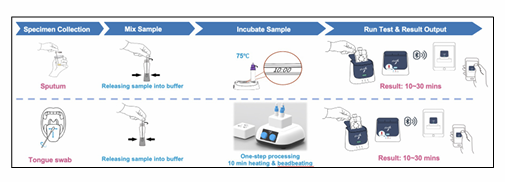

### 3. Supplemental Tables

#### Table S1. Classification for the reference standard for patients aged ≥15 years and corresponding numbers

| **TB case category** | | **Reference Standards** | | | | | **Total**  **(n=533)**  **n (%)** |
| --- | --- | --- | --- | --- | --- | --- | --- |
|  |  | **SMRS^a^: culture** | **EMRS^b^: culture or Xpert** | **EMRS: sensitivity analysis** | **CRS^c^: microbiological & clinical** | **CRS: sensitivity analysis** |  |
| **Definite TB** | |  |  |  |  |  | **134 (25.1)** |
| A1 | Culture positive MTB and Xpert positive (excluding trace) | TB | TB | TB | TB | TB | 105 (19.7)^d^ |
| A2 | Culture positive MTB and BD MAX positive | TB | TB | TB | TB | TB | 2 (0.4) |
| A3 | Culture positive MTB and Xpert positive trace | TB | TB | TB | TB | TB | 3 (0.6) |
| A4 | Culture positive MTB and Xpert negative | TB | TB | TB | TB | TB | 6 (1.1) |
| A5 | Culture positive NTM and Xpert positive (excluding trace) | Excluded | TB | TB | TB | TB | 1 (0.2) |
| A6 | Culture negative and Xpert positive (excluding trace) | Negative | TB | TB | TB | TB | 16 (3.0)^e^ |
| A7 | Culture negative and BD MAX positive | Negative | TB | TB | TB | TB | 1 (0.2) |
| A8 | Culture not done and Xpert positive (excluding trace) | Excluded | TB | TB | TB | TB | 0 (0.0) |
| **Probable TB (Culture or Xpert positive, Xpert trace for sensitivity analysis)** | |  |  |  |  |  | **73 (13.7)** |
| B1 | Culture negative and Xpert positive trace, CXR positive | Negative | Negative | TB | TB | TB | 5 (1.0) |
| B2 | Culture not done and Xpert positive trace, CXR positive | Excluded | Negative | TB | TB | TB | 0 (0.0) |
| B3 | Culture positive NTM and Xpert negative, and CXR positive, and treated for TB | Excluded | Negative | Negative | TB | TB | 4 (0.7) |
| B4 | Culture negative and Xpert negative and CXR positive and treated for TB | Negative | Negative | Negative | TB | TB | 63 (11.8) |
| B5 | Culture negative and BD MAX negative and CXR positive and treated for TB | Negative | Negative | Negative | TB | TB | 1 (0.2) |
| B6 | Culture negative, Xpert not done, CXR positive, and treated for TB | Negative | Negative | Negative | TB | TB | 0 (0.0) |
| B7 | Culture not done, Xpert negative, CXR positive, and treated for TB | Excluded | Negative | Negative | TB | TB | 0 (0.0) |
| B8 | Culture positive NTM, Xpert not done, CXR positive, and treated for TB | Excluded | Negative | Negative | TB | TB | 0 (0.0) |
| **Possible TB** | |  |  |  |  |  | **106 (19.9)** |
| C1 | Culture negative and Xpert positive trace, CXR negative, treatment unknown | Negative | Negative | TB | Negative | TB | 0 (0.0) |
| C2 | Culture negative, Xpert positive trace, CXR negative, treated for TB | Negative | Negative | TB | Negative | TB | 4 (0.7) |
| C3 | Culture not done and Xpert positive trace, CXR negative, treatment unknown | Excluded | Negative | TB | Negative | TB | 0 (0.0) |
| C4 | Culture negative and Xpert negative and CXR negative but treated for TB | Negative | Negative | Negative | Negative | Negative | 0 (0.0) |
|  |  | Negative | Negative | Negative | Negative | Excluded if EPTB^f^. | 8 (1.5) |
| C5 | Culture positive NTM and Xpert negative and CXR negative and not treated | Excluded | Negative | Negative | Negative | Negative | 6 (1.1) |
| C6 | Culture negative, Xpert not done, CXR negative, and treated for TB | Negative | Negative | Negative | Negative | Negative | 1 (0.2)^g^ |
| C7 | Culture positive NTM and Xpert negative, and CXR positive and not treated for TB | Excluded | Negative | Negative | Negative | Negative | 2 (0.4) |
| C8 | Culture negative and Xpert negative and CXR positive and not treated for TB | Negative | Negative | Negative | Negative | Negative | 79 (14.8) |
| C9 | Culture negative and BD MAX negative and CXR positive and not treated for TB | Negative | Negative | Negative | Negative | Negative | 6 (1.1) |
| C10 | Culture negative, Xpert not done, CXR positive, not treated for TB | Negative | Negative | Negative | Negative | Negative | 0 (0.0) |
| **Not TB (microbiological and clinical)** | |  |  |  |  |  | **220 (41.3)** |
| D1^h^ | Culture negative and Xpert negative and CXR negative and not treated for TB | Negative | Negative | Negative | Negative | Negative | 201 (37.7) |
| D2^h^ | Culture negative and BD MAX negative and CXR negative and not treated for TB | Negative | Negative | Negative | Negative | Negative | 7 (1.3) |
| D3^h^ | Culture negative, Xpert not done, CXR negative, and not treated for TB | Negative | Negative | Negative | Negative | Negative | 6 (1.1) |
| D4^h^ | Culture negative, Xpert negative, CXR not done, not treated for TB | Negative | Negative | Negative | Negative | Negative | 5 (0.9) |
| D5^h^ | Culture negative, BD MAX negative, CXR not done, not treated for TB | Negative | Negative | Negative | Negative | Negative | 1 (0.2) |
| D6^h^ | Culture not done, Xpert negative, CXR negative, and not treated for TB | Excluded | Negative | Negative | Negative | Negative | 0 (0.0) |
| D7 | Culture negative, Xpert not done, CXR not done, not treated for TB | Negative | Negative | Negative | Negative | Negative | 0 (0.0) |

^a^SMRS = Strict microbiological reference standard – Culture; ^b^EMRS = Microbiological reference standard – Culture or Xpert; ^c^CRS = Composite reference standard; ^d^Including seven patients with Xpert Ultra positive (five MTB detected, Rif resistance not detected and two MTB detected, Rif resistance detected) but not recorded semi-quantitative results; ^e^Including three patients with Xpert Ultra positive (2 MTB detected, Rif resistance not detected and 1 MTB detected, Rif resistance detected) but not recorded semi-quantitative results; ^f^EPTB = Extra Pulmonary TB; ^g^Participant was treated for TB based on a TST induration of 23 mm; ^h^Included for the formal calculation of test specificity.

#### Table S2: Demographic, clinical characteristics and laboratory findings of patients by site

| **Characteristics** | **Lung hospital**  **(n=192)**  **n (%)** | **Lung clinic**  **(n=335)**  **n (%)** | **CHC**  **(n=144)**  **n (%)** | **Total**  **(n=671)**  **n (%)** |
| --- | --- | --- | --- | --- |
| Female | 59 (30.7) | 164 (48.9) | 74 (51.4) | 297 (44.3) |
| Hospitalisation at enrolment | 131 (68.2) | 8 (2.4) | 0 (0.0) | 139 (20.7) |
| Aged 0 – 14 years | 16 (8.3) | 113 (33.7) | 9 (6.3) | 138 (20.6) |
| CXR |  |  |  |  |
| TB-specific abnormality | 143 (74.5) | 203 (60.6) | 42 (29.2) | 388 (57.8) |
| Non-TB abnormality | 35 (18.2) | 118 (35.2) | 42 (29.2) | 195 (29.1) |
| Normal | 13 (16.8) | 12 (3.6) | 53 (36.8) | 78 (11.6) |
| Not readable | 0 (0.0) | 1 (0.3) | 0 (0.0) | 1 (0.1) |
| Not done | 1 (0.5) | 2 (0.6) | 7 (4.9) | 10 (1.5) |
| Xpert Ultra result (n=586) |  |  |  |  |
| MTB detected | 81 (43.1) | 32 (12.2) | 25 (18.5) | 138 (23.6) |
| MTB not detected | 107 (56.9) | 231 (87.8) | 110 (81.5) | 448 (76.5) |
| Xpert Ultra semi-quantitative result (n=138) |  |  |  |  |
| High | 36 (44.4) | 4 (12.5) | 2 (8.0) | 10 (7.2) |
| Medium | 9 (11.1) | 3 (9.4) | 2 (8.0) | 12 (8.7) |
| Low | 18 (22.2) | 18 (56.3) | 4 (16.0) | 11 (8.0) |
| Very low | 4 (4.9) | 3 (9.4) | 10 (40.0) | 46 (33.3) |
| Trace | 8 (9.9) | 2 (6.2) | 5 (20.0) | 17 (12.3) |
| Not recorded | 6 (7.4) | 2 (6.2) | 2 (8.0) | 42 (30.4) |
| BD MAX result |  |  |  |  |
| MTB detected | 0 (0.0) | 3 (17.7) | 0 (0.0) | 3 (16.7) |
| MTB not detected | 1 (100) | 14 (82.3) | 0 (0.0) | 15 (83.3) |
| Culture results |  |  |  |  |
| Positive | 76 (39.6) | 36 (10.7) | 20 (13.9) | 132 (19.7) |
| Negative | 116 (60.4) | 296 (88.4) | 124 (86.1) | 536 (79.9) |
| Not done | 0 (0.0) | 3 (0.9) | 0 (0.0) | 3 (0.4) |
| Culture positive identification (n=132) |  |  |  |  |
| MTB | 70 (92.1) | 30 (83.3) | 18 (90.0) | 118 (89.4) |
| NTM | 6 (7.9) | 6 (16.7) | 2 (10.0) | 14 (10.6) |

#### Table S3: Distribution of alternative diagnoses in patients aged ≥15 years

| **Diagnosis** | **N = 198^a^**  **n (%)** |
| --- | --- |
| Chronic lung disease | 83 (41.9) |
| Upper respiratory tract infection | 48 (24.2) |
| Lower respiratory tract infection | 45 (22.7) |
| Cardiovascular conditions | 6 (3.0) |
| Neoplasm | 6 (3.0) |
| TB infection | 5 (2.5) |
| Miscellanous | 5 (2.5) |

^a^Of 220 diagnosed as Not TB, 198 had a specific alternative diagnosis recorded

#### Table S4: Distribution of alternative diagnoses in patients aged 0 – 14 years

| **Diagnosis** | **N=55^a^**  **n (%)** |
| --- | --- |
| Lower respiratory tract infection | 17 (30.9) |
| Chronic airway disease | 15 (27.3) |
| Lymphadenopathy | 10 (18.2) |
| Upper respiratory tract infection | 4 (7.3) |
| TB infection | 4 (7.3) |
| Miscellaneous condition | 5 (9.1) |

^a^Of 74 diagnosed as Unlikely TB, 55 had a specific alternative diagnosis recorded

#### Table S5: Time difference between index test and reference test specimen collection

| **Interval between index and reference specimen collection (days)** | **N = 671**  **n (%)** |
| --- | --- |
| 0 | 623 (92.8) |
| 1 | 40 (6.0) |
| 2 | 4 (0.6) |
| 3 | 4 (0.6) |

#### Table S6: Number of participants included in diagnostic accuracy analysis by reference standard

| **Reference standard** | **Total**  **(n)** | **Excluded (n)** | **Included (n)** | **TB positive (n)** | **TB negative (n)** |
| --- | --- | --- | --- | --- | --- |
| **Aged ≥15 years** | | | | | |
| SMRS | 533 | 13^b^ | 520 | 116 | 404 |
| EMRS | 533 | 0 | 533 | 134 | 399 |
| EMRS (sensitivity analysis) | 533 | 0 | 533 | 143 | 390 |
| CRS | 533 | 0 | 533 | 197 | 336 |
| CRS (sensitivity analysis) | 533 | 8^c^ | 525 | 211 | 314 |
| **Aged 0 – 14 years** | | | | | |
| SMRS – C | 138 | 2^a^ | 136 | 2 | 134 |
| EMRS – C | 138 | 2^a^ | 136 | 4 | 132 |
| CRS – C | 138 | 2^a^ | 136 | 74 | 62 |

^a^Two of 138 patients aged 0 – 14 years were excluded (one unclassified; one MiniDock invalid/error); ^b^Excluding 13 NTM cases; ^c^Excluding 8 EPTB cases.

#### Tabel S7: STARD checklist

| **Section & Topic** | **No** | **Item** | **Reported section** |
| --- | --- | --- | --- |
| TITLE OR ABSTRACT |  |  |  |
|  | 1 | Identification as a study of diagnostic accuracy using at least one measure of accuracy (such as sensitivity, specificity, predictive values, or AUC) | Abstract |
| ABSTRACT |  |  |  |
|  | 2 | Structured summary of study design, methods, results, and conclusions (for specific guidance, see STARD for Abstracts) | Abstract |
| INTRODUCTION |  |  |  |
|  | 3 | Scientific and clinical background, including the intended use and clinical role of the index test | Introduction |
|  | 4 | Study objectives and hypotheses | Introduction |
| METHODS |  |  |  |
| *Study design* | 5 | Whether data collection was planned before the index test and reference standard were performed (prospective study) or after (retrospective study) | Study design |
| *Participants* | 6 | Eligibility criteria | Study procedure |
|  | 7 | On what basis potentially eligible participants were identified (such as symptoms, results from previous tests, inclusion in registry) | Study procedure |
|  | 8 | Where and when potentially eligible participants were identified (setting, location and dates) | Study design |
|  | 9 | Whether participants formed a consecutive, random or convenience series | Study design |
| *Test methods* | 10a | Index test, in sufficient detail to allow replication | Index test, supplemental methods M1 |
|  | 10b | Reference standard, in sufficient detail to allow replication | Reference tests |
|  | 11 | Rationale for choosing the reference standard (if alternatives exist) | Statistical analysis |
|  | 12a | Definition of and rationale for test positivity cut-offs or result categories of the index test, distinguishing pre-specified from exploratory | Index test |
|  | 12b | Definition of and rationale for test positivity cut-offs or result categories of the reference standard, distinguishing pre-specified from exploratory | Reference tests |
|  | 13a | Whether clinical information and reference standard results were available to the performers/readers of the index test | Test methods |
|  | 13b | Whether clinical information and index test results were available to the assessors of the reference standard | Test methods |
| *Analysis* | 14 | Methods for estimating or comparing measures of diagnostic accuracy | Statistical analysis; Figure 1 |
|  | 15 | How indeterminate index test or reference standard results were handled | Index test; Reference tests; Statistical analysis |
|  | 16 | How missing data on the index test and reference standard were handled | Statistical analysis |
|  | 17 | Any analyses of variability in diagnostic accuracy, distinguishing pre-specified from exploratory | Statistical analysis |
|  | 18 | Intended sample size and how it was determined | Sample size |
| RESULTS |  |  |  |
| *Participants* | 19 | Flow of participants, using a diagram | Figure 2 |
|  | 20 | Baseline demographic and clinical characteristics of participants | Table 1 |
|  | 21a | Distribution of severity of disease in those with the target condition | Table 1 |
|  | 21b | Distribution of alternative diagnoses in those without the target condition | Table S3; Table S4 |
|  | 22 | Time interval and any clinical interventions between index test and reference standard | Table S5 |
| *Test results* | 23 | Cross-tabulation of the index test results (or their distribution) by the results of the reference standard | Table 2, Figure 3 |
|  | 24 | Estimates of diagnostic accuracy and their precision (such as 95% confidence intervals) | Figure 3 |
|  | 25 | Any adverse events from performing the index test or the reference standard | N/A |
| DISCUSSION |  |  |  |
|  | 26 | Study limitations, including sources of potential bias, statistical uncertainty, and generalisability | Discussion |
|  | 27 | Implications for practice, including the intended use and clinical role of the index test | Discussion; Conclusion |
| OTHER INFORMATION |  |  |  |
|  | 28 | Registration number and name of registry |  |
|  | 29 | Where the full study protocol can be accessed | On request |
|  | 30 | Sources of funding and other support; role of funders | Abstract; Notes – Financial support |
